# Clinical implementation of a machine learning system to detect deteriorating patients reduces time to response and intervention

**DOI:** 10.1101/2021.10.10.21264823

**Authors:** Santiago Romero Brufau, Jacob Rosenthal, Jordan Kautz, Curtis Storlie, Kim Gaines, Jill Nagel, Adam VanDeusen, Matthew Johnson, Joel Hickman, Dale Hardin, Julie Schmidt, Gene Dankbar, Jeanne Huddleston

## Abstract

**Introduction:** Acute physiological deterioration is a major contributor to in-hospital morbidity and mortality. Early detection and intervention of deteriorating patients is key to improving patient outcomes. Prior research has demonstrated the effectiveness of Early Warning Systems and other algorithmic approaches in automatically identifying these patients from passively monitoring vital signs.

**Methods:** In this work, we conduct a prospective pilot study of clinical deployment of the Mayo Clinic Bedside Patient Rescue (BPR) system using an escalating alerting logic enabled by machine learning. Among four units where the BPR system was deployed, time to response and time to intervention for deteriorating patients were significantly reduced relative to matched control units.

**Results:** In pilot units, time to response decreased by 35.4% (from 63.2 minutes to 40.8 minutes) and time to intervention decreased by 48.5% (from 106.3 minutes to 55.9 minutes). No significant differences were observed in counterbalance metrics of mortality, ICU transfer rate, and Rapid Response Team activation rate. Furthermore, the automated alerting system was well-received by clinicians participating in the pilot study, as assessed by survey.

**Discussion:** These results demonstrate a successful clinical deployment of a practice-changing machine learning alert system with demonstrable impact on improving patient care.

## Introduction

Physiological deterioration, defined here as any significant worsening in the condition of a hospitalized patient that can result in patient morbidity or mortality, is a major cause of death in the hospital setting. Deterioration may be triggered by events such as sepsis or acute respiratory failure, or may emerge as a result or complication of care provided. Many instances of physiological deterioration of hospitalized patients result in death, cardiac arrest, or an increase in the intensity of care that is needed to support the patient’s physiology, resulting in an unplanned admission to the intensive care unit (ICU). The overall incidence of these events is estimated to be around 2% per patient-day^1^, and about 17 patients per 100 beds per year for in-hospital cardiac arrest^2^, of which only about 18% survive to discharge^2,3^.

Early intervention for physiological deterioration is a key determinant of patient outcomes. For example, delayed transfer of critically ill patients to the intensive care unit has been shown to be associated with increased mortality^4,5^. It is also known that, in the case of septic shock (one of the main causes of in-hospital deterioration), mortality increases approximately 8% for every hour that antibiotic treatment is delayed^6^. Additionally, the mortality rate for patients who develop sepsis in the hospital is significantly higher compared to patients that present with sepsis at the time of hospital admission, and most deaths related to sepsis come from patients presenting with a less severe form of sepsis^7^ whose condition later deteriorates. This suggests that timely detection of deterioration in hospitalized patients is a key contributor to preventable mortality.

One way to reduce intervention times, and thus improve patient outcomes, is to identify deteriorating patients as early as possible. Up to 85% of severe adverse events (such as cardiac arrest, death, or emergency intensive care admission) that happen in the hospital are preceded by abnormal vital signs^8-11^, suggesting that they may be identifiable by an algorithmic approach. This intuition has motivated development of several identification algorithms for the detection of acute deterioration, known as early warning scores or early warning systems (EWS)^12,13^, previously known as track-and-trigger scores or track-and-trigger systems^14,15^. In prior work, we employed machine learning (ML) to develop an EWS to predict inpatient deterioration (MC-EWS)^16^.

However, having a predictive model is of little use unless the prediction can be performed in real-time and it results in an early alert in real clinical situations in which a patient is deteriorating. Additionally, it is important to test whether an early alert results in an early intervention, and how much time can be saved. Here, we conduct a pilot study of implementation of this system in the clinic using a cascading alert protocol and phased implementation approach and evaluate its effect on time to intervention and time to therapy for physiological deterioration.

### Related work

Complex predictive models that require the use of EHR data have been developed for the prediction of acute inpatient deterioration or death^17-19^, including at least two that were tested in a randomized controlled trial where alerts were sent to a Rapid Response Team^20^ or were channeled through a centralized review team^21^. Other studies have implemented automated versions of simpler paper-based early warning scores^22^ such as the National Early Warning Score^23^, or automated vital signs triggers^24^. Results from these efforts have been mixed.

Typically, studies that have seen positive results have directed alerts at a centralized location or required a dedicated team to triage the alerts, which may not be feasible in all hospitals.

### Our approach

We developed an automated alert system based on the principle of “time-limited escalation of expertise to the bedside” that does not require review by a dedicated team, and then tested it in a controlled, pragmatic trial.

## Methods

### Setting and standard of care

This study was conducted at the Mayo Clinic, a tertiary academic medical center in the Midwest United States. The alerting system was implemented in 4 hospital units (2 general internal medicine units and 2 colorectal surgery units), totaling about 100 beds. The study was approved by the Institutional Review Board, the Hospital Practice Subcommittee, the Mayo Clinic Clinical Practice Committee, and many others.

In the study units, the standard of nursing care during the period of the study was to obtain a set of vitals every 8-12 hours. Vital signs are manually entered by nursing staff into the electronic medical record, or automatically collected by sensors and confirmed by a nurse. Nursing staff has the flexibility to increase the frequency of vital sign monitoring at their discretion. Vital signs on medical and surgical floors with telemetric capability are collected more frequently with electronic capture and recorded into the electronic medical record. Laboratory studies are ordered at the physician discretion at any time of day. The highest volume of laboratory orders occurs early in the morning so that results are available for morning physician team rounds. The Rapid Response Team (RRT) responds at the request of nurses or physicians in both the general care and telemetry floors of the hospitals. The RRT completed rollout in 2006 so it is a well-established part of the workflow. The RRT is a multidisciplinary team including: respiratory therapist, critical care nurse, and either a critical care fellow or an intensivist. Institutional guidelines suggest RRT activation when a patient has a new and/or unexpected change with any of several criteria (**Error! Reference** s**ource not found**.).

### Machine learning model

The Mayo Clinic Early Warning Score (MC-EWS) has been described in detail previously^16^. Briefly, it comprises a gradient boosting model (GBM) trained to predict occurrence of any of three deterioration events: transfer to intensive care unit (ICU), resuscitation call, or RRT team activation. The model was trained on two years of historical data, including raw clinical data and engineered features (e.g., time series information). Model performance was assessed on a held-out validation set from the same hospital system, and also externally validated using data from a tertiary care hospital in the Southwest United States. For this implementation study, we combined the MC-EWS score and the nursing worry factor (WF) score using a linear model to create the Bedside Patient Rescue (BPR) score:

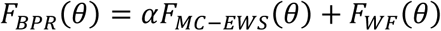

Where θ represents the clinical data, *F*_*MC™EWS*_*(θ)* represents the machine learning MC-EWS score, *F*_*WF*_ *(θ)* represents the nursing WF score, and *α* is a parameter used to weight the two scores to compute the final BPR score.

### Technical Implementation

For implementation of the BPR scoring system in the clinic, we designed an automated workflow to calculate the BPR score for each patient at regular 15-minute intervals. The system is directly integrated with the electronic health record (EHR) and fetches the relevant clinical data from the EHR real-time database whenever a score is calculated. We manually created mappings between the features of interest and the items in the EHR data schema to ensure consistency and prioritize the most recent data. For example, if heart rate has been measured peripherally by a clinician and also electronically from EKG, those will be different entries in the database, so logic is implemented to select the most recent data point before feeding it as input to the model. Upon score calculation, if the score is above the threshold, the cascading alert logic is triggered automatically (Figure 1).

**Figure 1.**
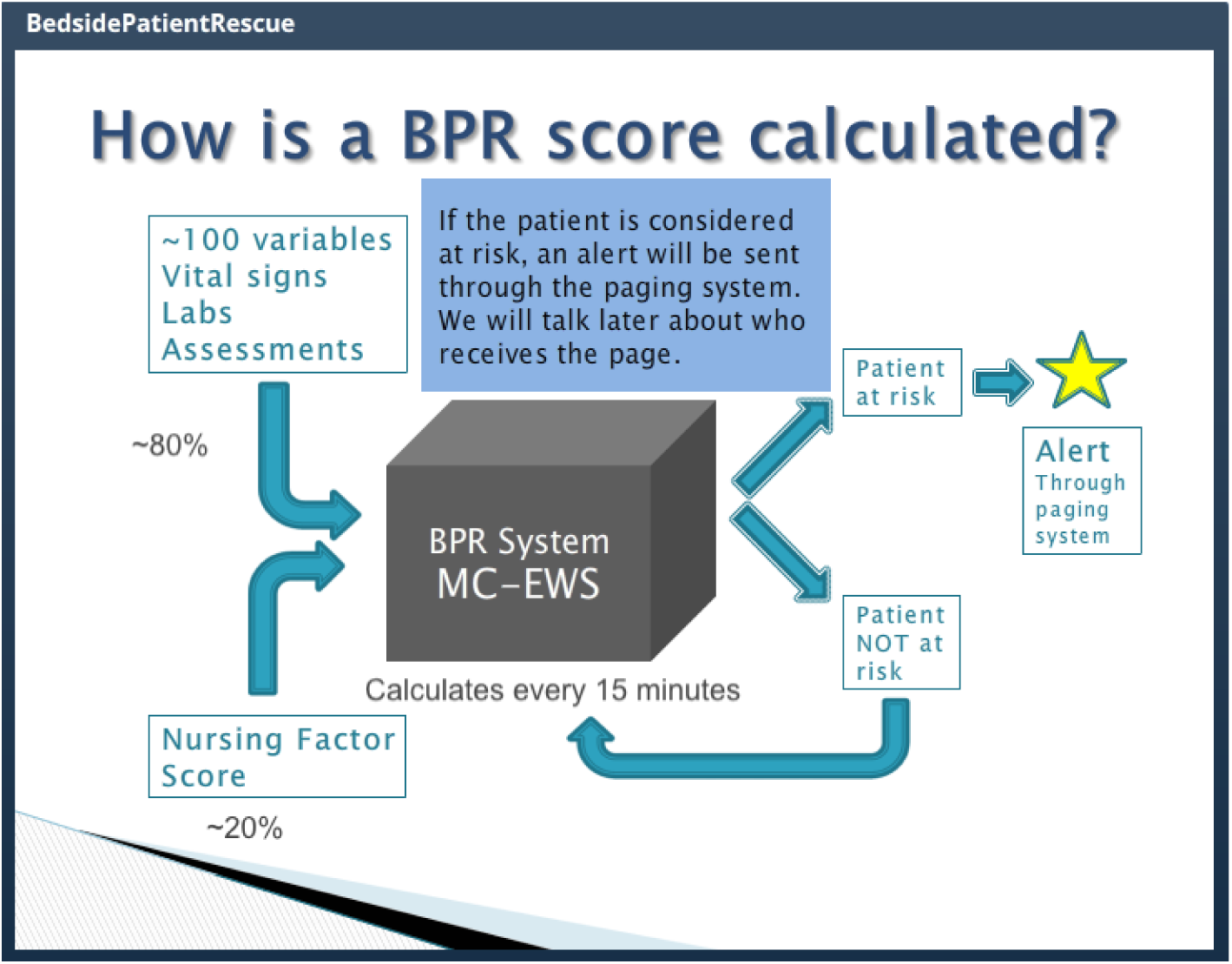
Schematic of BPR system

### Escalating Alert Logic

The automated alerting is designed as a human-in-the loop system based on the principle of “time-limited escalation of expertise to the bedside.” Upon trigger by a BPR score above the specified threshold, a cascading sequence with four distinct phases is initiated (Figure 2). The threshold was chosen based on simulated alert frequency on retrospective dataset and tuned during the silent deployment phase. This design ensures that clinicians are promptly alerted to patients suspected to be at high risk of deterioration, while minimizing the risk of inducing alert fatigue.

**Figure 2.**
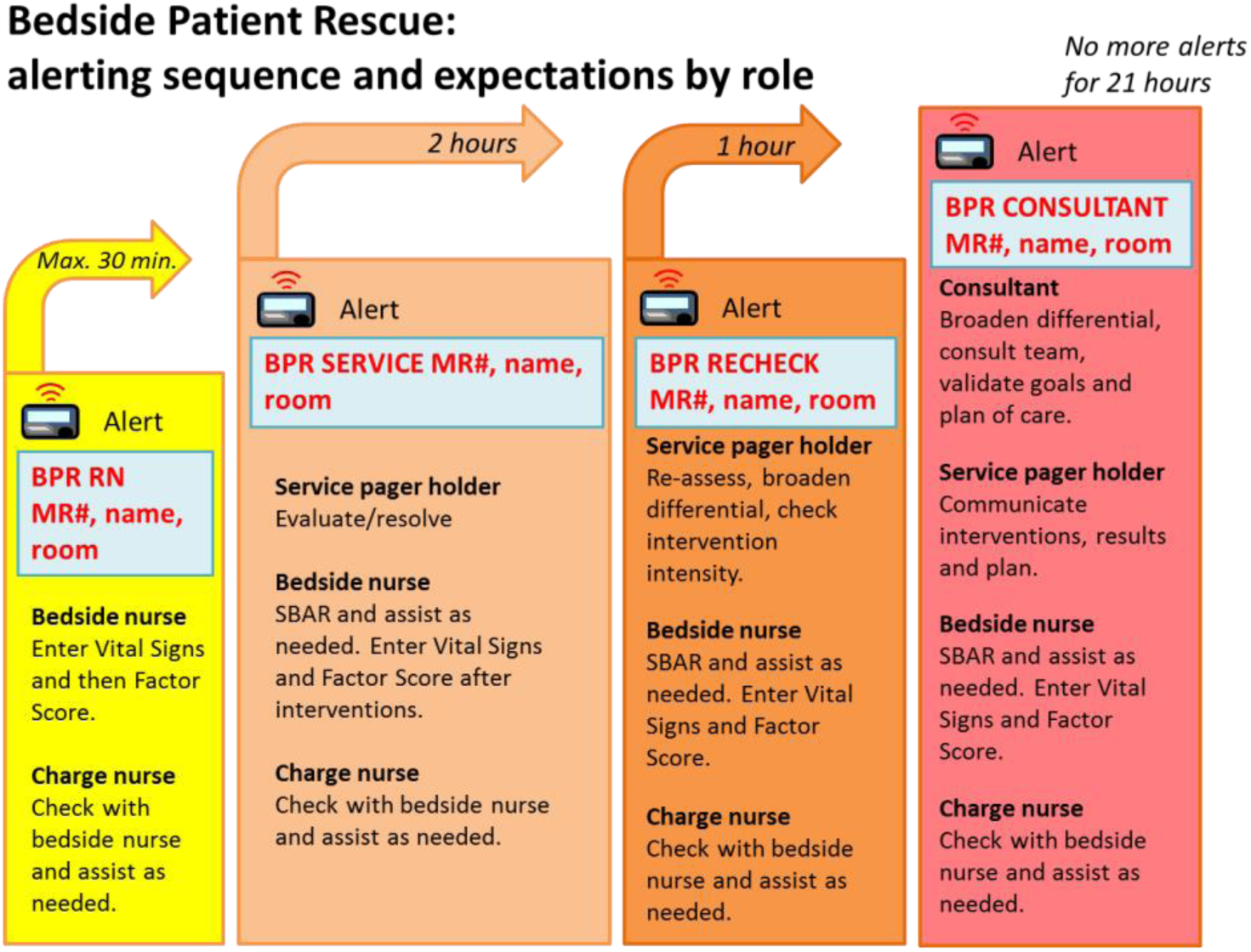
Escalating alert logic of the BPR scoring system

The first alert is triggered when the BPR score is above the threshold. At that point, a notice is sent to the pagers of the nurse assigned to the patient, as well as the charge nurse for the unit. The nursing team is prompted to check on the patient, update vital signs into the EMR, and enter an updated worry factor score within 30 minutes of the alert. Immediately upon entry of the updated clinical data, the BPR score is automatically recalculated. If the recalculated BPR score is still above the threshold, then the alert is automatically escalated.

In the second phase of the escalation protocol, an alert is sent to the service pager for the unit, which is typically monitored by a resident physician, physician assistant, or nurse practitioner. At that point, the service pager holder must evaluate the patient within 2 hours. That person is responsible for initiating an intervention if necessary. After intervention, or at the end of the 2-hour window, vital signs are again updated and a new WF score is assessed by the bedside nurse. The BPR score is then recalculated and automatically evaluated for further escalation.

The third escalation also goes to the service pager for the unit. The care team is prompted to re-evaluate the patient, considering broader differential diagnoses and checking the intensity of any prior intervention from the previous phase. Again, vital signs and WF are updated and the BPR score is recalculated automatically at the end of the time window, which in this phase is restricted to one hour. If the score is still above the threshold, the alert is escalated once more.

The fourth and highest escalation level triggers an automatic alert to the attending physician for the patient. At this point, the entire care team works together to evaluate the patient and determine a plan of care. This fourth phase is the final step in the cascading protocol; at this point, the entire care team is involved and aware of the situation, so no further alerting is necessary. All further alerts for the patient were therefore disabled for 21 hours after escalation to the fourth phase, thus restricting the entire escalation protocol to only happen once in a 24hr window.

The cascading alert protocol defines the baseline for escalation, but at any step in the process members of the care team are able to manually escalate based on their judgement. For example, after the first alert, if the bedside nurse believes that the patient is at significant risk and it is appropriate to call a Rapid Response Team, it is encouraged that he/she do so.

Additional filters were applied in the implementation of the alert flow logic to account for particular situations. For example, patients in hospice care or “comfort care only” designation were excluded from all alerts, as were patients in the operating room. At every escalation in the alerting cascade, all members of the care team who had been previously alerted were alerted again, to keep them in the loop. The escalating alert logic described in the previous paragraphs was developed in consultation with patient care team members, including which care team members are alerted at each level and the duration of time between alerts.

### Phased implementation approach

In this study, the BPR scoring and alert escalation system was implemented in 4 hospital units, totaling about 100 beds, with each unit in the intervention matched with a control unit similar in clinical practice and patient population. In all eight units (intervention and control), the nursing worry factor was routinely entered once per patient per nursing shift. The system was initially run silently for 4 months in all eight units before the pilot was started, to gather baseline data. During this silent phase, BPR scores were calculated but alerts were routed to a database instead of being sent to pagers on the care team. The study team analyzed the alerting data from the silent deployment and calibrated the alerting threshold for BPR score to ensure that the average frequency of alerts would not exceed a pre-specified threshold of 1 alert per day per 10 patients, to reduce risk of alert fatigue. After the silent deployment phase, the system went live in the four intervention units on a staggered schedule (Figure 3). The date when the system was activated defined the beginning of the post-treatment phase in each unit and its control.

**Figure 3.**
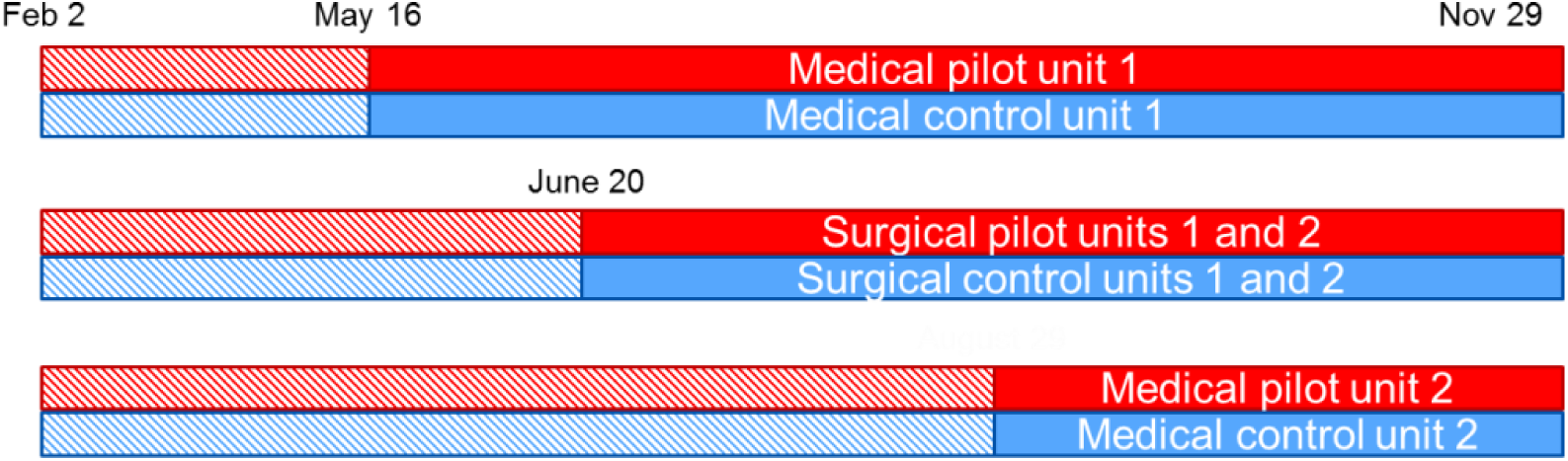
Phased implementation of the BPR pilot

### Coordination with staff before implementation

Coordination with clinical staff was prioritized at all phases of this study, starting from the design phase, when user testing of the alert logic and the wording of the alerts was performed in Mayo Clinic’s simulation center using simulated patients and a volunteer team of nurses, resident physicians and attending physicians from both a medical and a surgical unit. To prepare the participating units for the significant change in workflow that implementation of the BPR system would entail, the study team facilitated a series of workshops for the clinical staff and distributed informational materials. The ADKAR change management framework^25^ was used to ensure that all members of the care team were willing and able to be active participants in the trial. Clinician buy-in is important for any modification to clinical workflows, but especially so in the context of the BPR score due to the integration of nursing worry factor scores.

### Outcome metrics

The primary outcomes of interest were time to intervention and time to response. Time to intervention was defined as the interval between onset of patient deterioration and any order placed for that patient (laboratory tests, medication, radiology exams, etc.). Time to response was defined as the interval between onset of patient deterioration and administration of medication, fluids, or supplemental oxygen. These metrics were measured in the pre-pilot period and post-intervention period, and units in the intervention group and their corresponding control units were compared using a difference-in-differences analysis^26^.

In addition to the primary metrics, we also tracked several counterbalance metrics to assess for adverse effects arising from implementation of the BPR system: number of RRT calls per 100 patient-days, ICU transfers per 100 patient-days, and mortality per 100 patient-days. We compared these in both pilot and control units during pre- and post-intervention periods. All differences were tested using the “N-1” Chi-squared test.

Finally, upon completion of the pilot study, we administered an online survey to all participating clinical staff to gauge providers’ experiences and impressions of the changes to the workflow.

## Results

Our cohort comprised a total of 11,665 hospitalizations, accounting for 54,825 cumulative days of hospitalization (Table 1).

**Table 1.** Cohort demographics

|  | Pre-Intervention |  | Post-Intervention |  |  |
| --- | --- | --- | --- | --- | --- |
|  | Control | Pilot | Control | Pilot | Total |
| **N** | 2440 | 3603 | 2705 | 2917 | 11665 |
| **Age at Admission** |  |  |  |  |  |
| Mean (SD) | 61.6 (20.07) | 60.4 (18.28) | 62.1 (18.60) | 59.8 (18.16) | 60.9 (18.73) |
| **Patient Gender, n (%)** |  |  |  |  |  |
| Female | 1230<br>(50.41%) | 1820<br>(50.51%) | 1421<br>(52.53%) | 1491<br>(51.11%) | 5962<br>(51.1%) |
| Male | 1210<br>(49.59%) | 1783<br>(49.49%) | 1284<br>(47.47%) | 1426<br>(48.89%) | 5703<br>(48.9%) |
| **Race, n (%)** |  |  |  |  |  |
| American Indian/Alaskan Native | 9 (0.4%) | 24 (0.7%) | 21 (0.8%) | 22 (0.8%) | 76 (0.7%) |
| Black | 63 (2.6%) | 116 (3.2%) | 66 (2.4%) | 73 (2.5%) | 318 (2.7%) |
| White | 2208<br>(90.5%) | 3255<br>(90.3%) | 2447<br>(90.5%) | 2642<br>(90.6%) | 10552<br>(90.5%) |
| Hawaiian/Pacific Islander | 4 (0.2%) | 1 (0.0%) | 0 (0.0%) | 4 (0.14%) | 9 (0.1%) |
| Other | 79 (3.2%) | 138 (3.8%) | 91 (3.4%) | 96 (3.3%) | 404 (3.5%) |
| Asian | 53 (2.2%) | 48 (1.3%) | 48 (1.8%) | 36 (1.2%) | 185 (1.6%) |
| Choose Not to Disclose/Unknown | 24 (1.0%) | 21 (0.6%) | 32 (1.2%) | 44 (1.5%) | 121 (1.0%) |
| **Length of Stay** |  |  |  |  |  |
| Mean (SD) | 4.7 (7.15) | 4.4 (5.88) | 4.6 (7.47) | 5.1 (8.37) | 4.7 (7.21) |

### Primary outcomes

Median time to response decreased by 35.4% (22.38 minutes) among the four intervention units, a significant (p<0.001) reduction compared to the control units, where time to response increased modestly by 6.5% (5.52 minutes). Similarly, median time to intervention decreased by 47.4% (50.46 minutes) among the four intervention units, representing a significant (p<0.001) reduction compared to the control units, where time to intervention increased by 12.4% (13.62 minutes) (Figure 4). Notably, the effect of intervention is driven by the surgical units, as no significant difference was observed in the medical units between intervention and control groups (**Error! Reference source not found**.).

**Figure 4.**
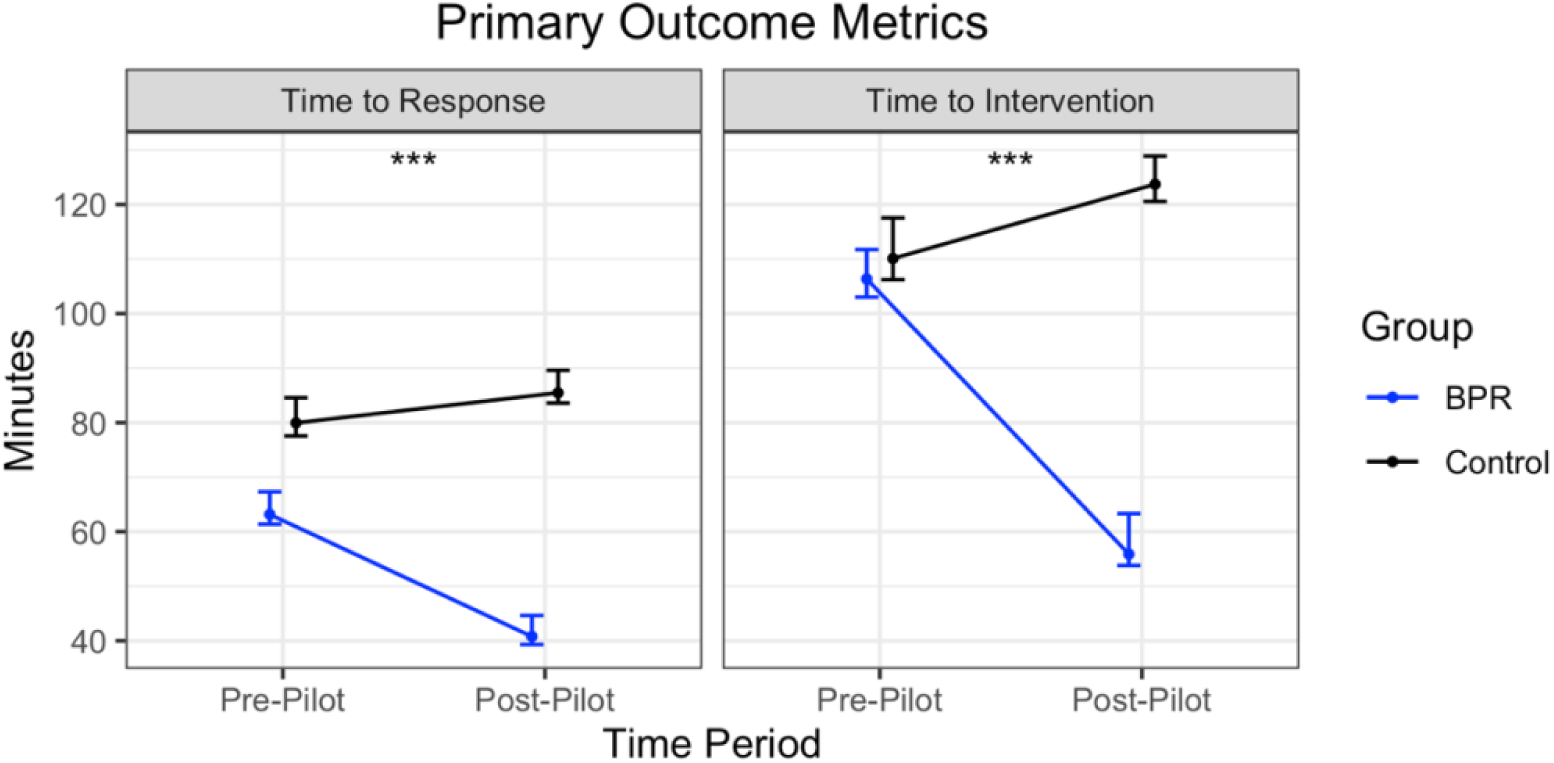
Primary outcome metrics

### Counterbalance metrics

We observed no significant change in any of the counterbalance metrics tracked in this study (Figure 5). In contrast with the primary outcome metrics, the counterbalance metrics showed no difference between medical and surgical units (**Error! Reference source not found**.). Due to low baseline rates of these metrics (baseline mortality is around 1%, baseline ICU transfer rate around 3%, and baseline rate of calls to the Rapid Response Team around 4%), we may have lacked power to detect subtle changes. However, we can say that implementation of the BPR system was not associated with any large-magnitude changes.

**Figure 5.**
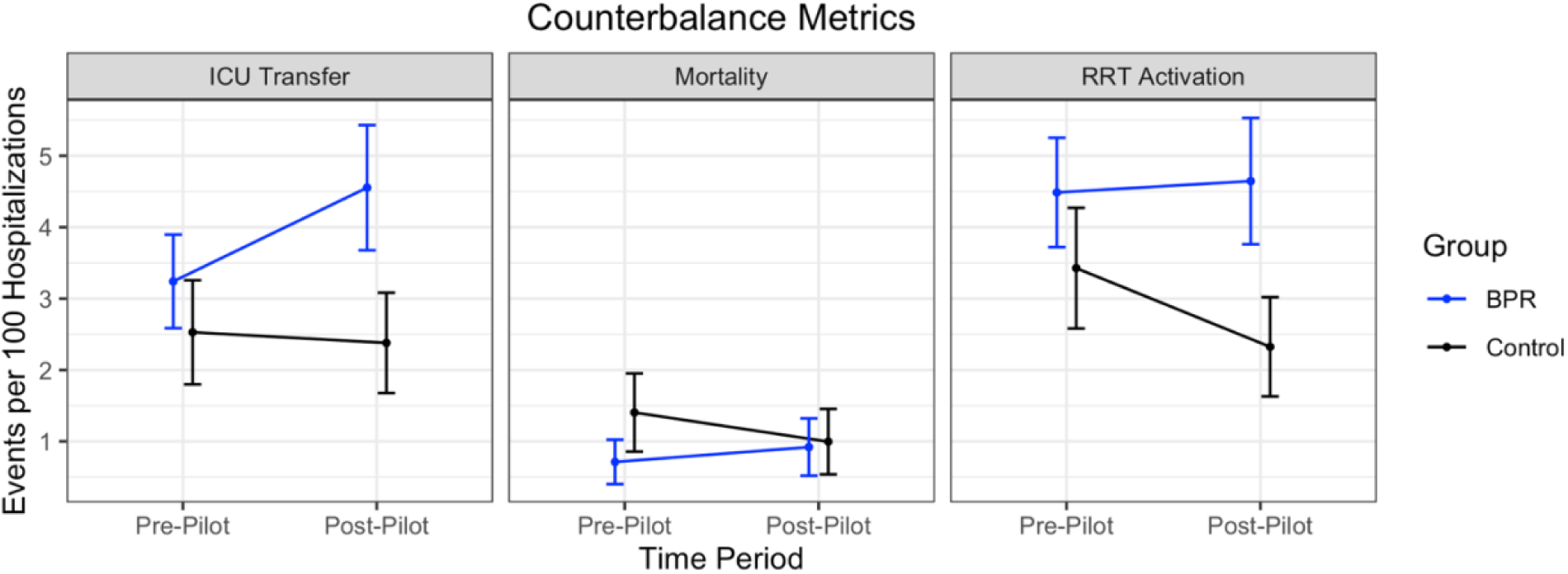
Counterbalance metrics

### Clinician acceptance

We received 63 responses out of a total of approximately 120 online surveys sent to the clinical staff in participating units. 59% of respondents agreed or strongly agreed that “Overall, I think the BPR project improves patient safety and I would like to see it continue.” Additionally, 81% agreed or strongly agreed to the statement “Alerts would be useful for team members who are less experienced.” (**Error! Reference source not found.**). These results suggest that despite the increase in workload, clinicians were enthusiastic about the integration of ML-powered alerting systems to improve patient outcomes. These also highlight the value of careful integration with existing workflows, and collaboration with staff at all phases of the implementation process.

## Discussion

In this study, we piloted clinical implementation of the Mayo Clinic Bedside Patient Rescue (BPR) score, a machine learning-powered automated alerting system for the detection of acute physiological deterioration. In all four units where the system was piloted, we observed a significant decrease in the primary metrics of time to response and time to intervention relative to matched control units. We observed no adverse effects, as measured by three counterbalance metrics. To our knowledge, this represents the first implementation study of its kind to demonstrate practice-changing integration of machine learning into the clinical setting with measurable improvements in patient care.

This study also provides a blueprint for further design and implementation of “human-in-the-loop” ML systems in the medical setting. At every stage in the process, we sought to integrate the system thoughtfully into existing clinical workflows, rather than supplant them. Integration of the nursing worry factor score into the final ensemble model was one means by which the system incorporated human inputs; equally important was the close collaboration with clinical staff at all phases of the study, from designing the cascading alerting logic to rolling it out in the clinic. The resulting implementation of the BPR system changed practice and improved patient care without disrupting clinical workflows or inducing alert fatigue. In fact, despite the increased workload from responding to alerts and entering the worry factor, the system received favorable impressions from participating clinical staff.

Although we saw a significant reduction in our primary outcome metrics in all four implementation units, the medical control units also saw a significant reduction. This could be due to cross-contamination and an increased focus on deterioration by clinicians, as some staff are shared between the medical control and intervention units, which is less the case in the surgical units. Further studies will be needed to further ascertain this effect.

These results may not generalize to all other settings with different practices. Nursing expertise is a direct input into the model (worry factor score), so results may vary in settings where the practices for nurse education differ. Furthermore, the BPR alerting system depends on buy-in from clinicians. After all, an alert is ultimately useless if it does not result in a change in actions by clinicians. Adoption may be different in settings with different cultures. Although we tested in 4 separate units, they are all in the Mayo Clinic system so these experiences may not generalize to cultures in other systems. We identified the education and implementation process as being critical to achieving good uptake by clinicians. We hope that studies such as this one which show the improvements to patient outcomes will help drive adoption.

Future work should address whether the demonstrated reduction in response time results in a reduction in mortality. Our 6-month pilot was not powered to detect smaller changes in mortality rate given the very low incidence of acute deterioration, and the even lower mortality rate in our hospital units. A longer pilot, or a pilot in an institution with a higher baseline mortality could accomplish that. Further studies should also explore model interpretability for clinicians, for example alerting the clinical staff to specific vital signs contributing a high BPR score. Greater transparency of alerts may increase trust of clinicians. Finally, we believe that alerting systems should engage user interface/user experience professionals to design the interfaces by which clinicians interact with the alerting system, with a specific focus on minimizing alert fatigue and friction with integration into workflows.

## Data Availability

Data produced in the present study are available upon reasonable request to the authors.

