## Supplemental Materials for "Clinical implementation of a machine learning system to detect deteriorating patients reduces time to response and intervention"

*Supplemental Table 1 Criteria for activation of RRT. Acute is defined as new and/or unexpected*

| |
|---|
| A staff member is worried about the patient |
| Acute and persistent declining oxygen saturations <90% |
| Acute and persistent change in heart rate: <40 beats/min or >130 beats/min |
| Acute and persistent change in systolic blood pressure: <90 mm Hg |
| Acute and persistent change in respiratory rate: <10 breaths/min or >28 breaths/min |
| Acute chest pain suggestive of ischemia |
| Acute and persistent change in conscious state (including agitated delirium) |
| New onset of symptoms suggestive of stroke |

*Supplemental Table 2 Primary outcome metrics*

| Metric | Group | Pre- intervention (95% CI) | Post-intervention (95% CI) | Difference (percentage change) | Difference in Differences |
|---|---|---|---|---|---|
| Time to Response | Pilot | 63.17 (61.39 - 67.32) | 40.79 (39.31 - 44.63) | -22.38 (-35.4%) | 27.90 (P<0.001) |
| | Control | 79.95 (77.55 - 84.57) | 85.47 (83.60 - 89.59) | 5.52 (6.5%) | |
| Time to Intervention | Pilot | 106.34 (103.02 - 111.74) | 55.88 (53.80 - 63.32) | -50.46 (-47.4%) | 64.08 (P<0.001) |
| | Control | 110.10 (106.23 - 117.54) | 123.72 (120.56 - 128.88) | 13.62 (12.4%) | |

*Supplemental Table 3 Counterbalance metrics*

| Metric | Group | Pre-intervention (95% CI) | Post-intervention (95% CI) | Difference | Difference in Differences |
|---|---|---|---|---|---|
| Mortality rate (deaths per 100 hospitalizations) | Pilot | 0.71 ± 0.31 | 0.92 ± 0.4 | 0.21 | 0.61 (P>0.05) |
| | Control | 1.40 ± 0.55 | 1.00 ± 0.46 | -0.40 | |
| ICU transfer rate (ICU transfers per 100 hospitalizations) | Pilot | 3.24 ± 0.65 | 4.55 ± 0.88 | 1.31 | 1.46 (P>0.05) |
| | Control | 2.53 ± 0.73 | 2.38 ± 0.7 | -0.15 | |
| RRT activation rate (RRT calls per 100 hospitalizations) | Pilot | 4.49 ± 0.77 | 4.65 ± 0.88 | 0.16 | 1.27 (P>0.05) |
| | Control | 3.43 ± 0.85 | 2.32 ± 0.69 | -1.11 | |

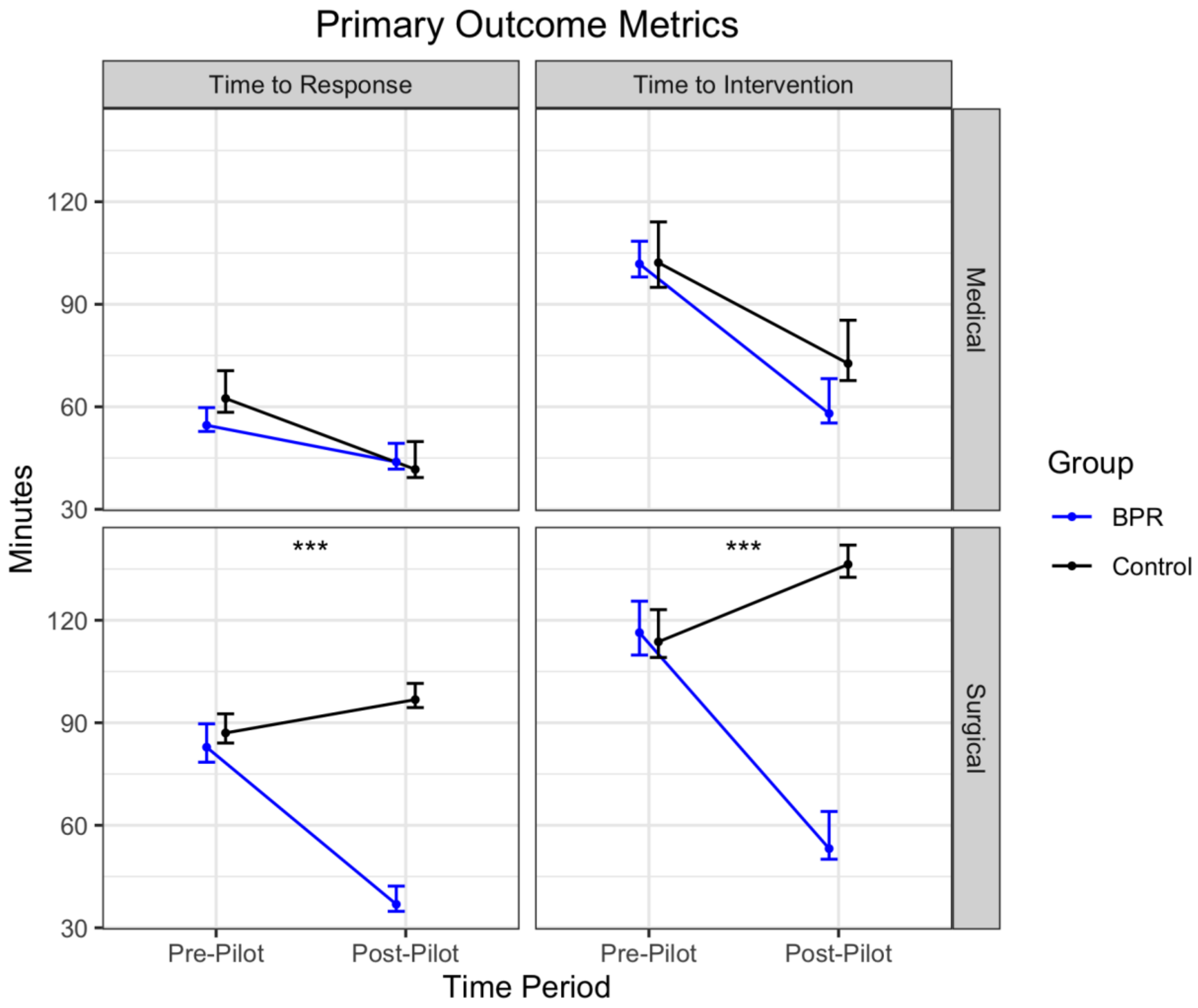


*Supplemental Figure 1 Primary outcome metrics by unit type*

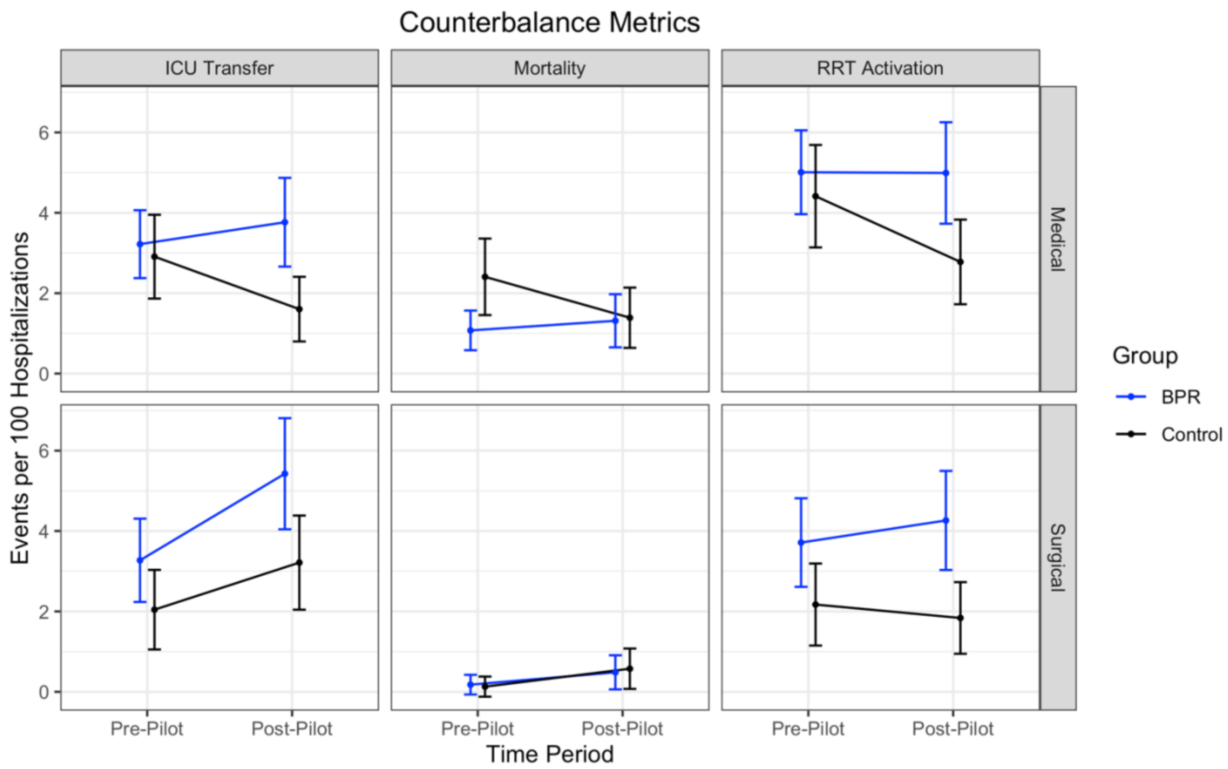


*Supplemental Figure 2 Counterbalance metrics by unit type*

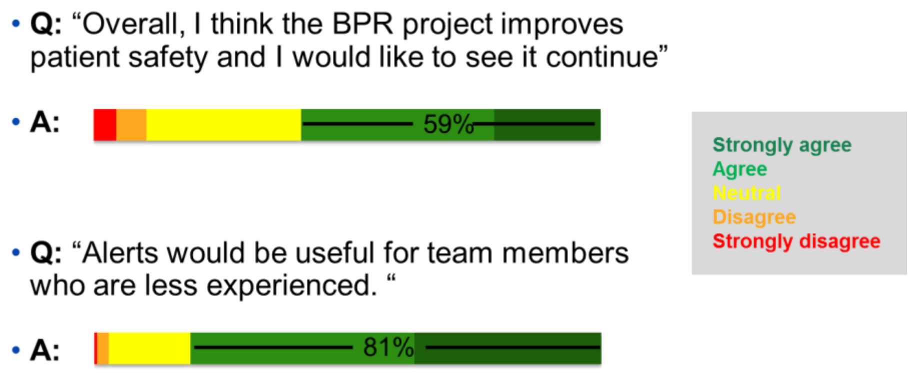


*Supplemental Figure 3 Survey results from participating clinicians*
